# Expanding clinical and variant spectrum of *CBX1*-related syndrome: Report of three novel cases

**DOI:** 10.64898/2026.09.28.26363820

**Authors:** Graciela Carrillo, Shuhei Fujino, Harrison Higgs, Anita Kabahizi, Randy Le, Rachel Rabin, John Pappas, Amy Crunk, Kara Ranguin, Jérémie Mortreux, AURAGEN consortium, Marilyn Lackmy, Carlos A Gadia, Matthew Hoi Kin Chau, Jill A. Rosenfeld, Xi Luo, Kosuke Izumi

**Affiliations:** Division of Genetics and Metabolism, Department of Pediatrics, University of Texas Southwestern Medical Center and Children’s Medical Center Dallas, Dallas, TX, USA; Clinical Genetic Services, New York University Grossman School of Medicine, New York, NY, USA; GeneDx, LLC, Gaithersburg, MD, USA; Clinical Genetics Unit, Guadeloupe University Hospital; GCS AURAGEN, Lyon, France; Nicklaus Children’s Hospital, Miami, FL, USA; Department of Obstetrics and Gynaecology, The Chinese University of Hong Kong, Hong Kong SAR, China; The Chinese University of Hong Kong-Baylor College of Medicine Joint Center for Medical Genetics, Hong Kong SAR, China; Department of Molecular and Human Genetics, Baylor College of Medicine, Houston, TX, USA; Baylor Genetics, Houston, TX, USA

## Abstract

*CBX1*-related syndrome is characterized by developmental delay, hypotonia, autistic features, and mild dysmorphic features. This syndrome is caused by heterozygous missense variants in *CBX1*, which encodes heterochromatin protein 1 beta (HP1β). These variants are situated within the chromodomain of HP1β, a critical region that mediates the interaction between HP1β and trimethylated H3K9. While a prior study identified three individuals with pathogenic *CBX1* variants, the variant and clinical spectrum of *CBX1*-related syndrome remains largely unknown. Here we report three patients with novel *de novo* heterozygous variants in *CBX1*. Patients with *CBX1* p.(Leu40Pro) and p.(Cys60Arg) manifested with global developmental delay, while the patient with *CBX1* p.(Cys60Gly) primarily exhibited speech delay without concurrent motor developmental delay. All variants reside within chromodomain of the HP1β protein. Interestingly, *in vitro* cellular assays revealed that *CBX1* p.(Cys60Gly) variant showed an intermediate cellular phenotype between the wild type and the *CBX1* p.(Leu40Pro) and p.(Cys60Arg) variants, suggesting a possible genotype-phenotype correlation. This report provides further evidence that missense variants within the chromodomain of HP1β cause a syndromic neurodevelopmental disorder, *CBX1*-related syndrome.

## Introduction

*CBX1*-related syndrome is characterized by developmental delay, hypotonia, autistic features, and mild dysmorphic features^1^. This syndrome is caused by heterozygous missense variants in *CBX1*, which encodes heterochromatin protein 1 beta (HP1β), a core structural protein of heterochromatin^2^. A prior study identified three individuals with pathogenic *CBX1* variants, c.151A>C; p.(Thr51Pro), c.155G>T; p.(Trp52Leu), and c.169A>G; p.(Asn57Asp). These variants are situated within the functional chromodomain (CD) of HP1β, a critical region that mediates the interaction between HP1β and trimethylated H3K9 (H3K9me3)^2,3,4^. *In vitro* cellular assays have shown that these mutant HP1β proteins lack the ability to bind heterochromatin, suggesting that *CBX1*-related syndrome results from the reduced binding of HP1β to H3K9me3-marked chromatin^1^. Although previous research has established a link between *CBX1* variants and neurodevelopmental disorders, significant gaps in knowledge remain, including a limited understanding of the phenotypic and variant spectra of *CBX1*-related syndromes, which pose a diagnostic challenge in clinical practice. The purpose of this article is to report three additional cases of *CBX1*-related syndrome, thereby contributing to a broader understanding of the phenotypic and mutational spectrum associated with this neurodevelopmental disorder.

## Material and Methods

### Ethical approval

This study was determined to be exempt from Institutional Review Board (IRB) review because only de-identified information was collected (University of Texas Southwestern Medical Center IRB number: STU20261615). Written informed consent was waived, as no identifiable information was obtained. Verbal informed consent was obtained from the patients’ families for the use of clinical data.

### *CBX1* variant computational prediction

In silico pathogenicity prediction was performed by using Franklin (https://franklin.genoox.com/clinical-db/home) and MutationTaster^5^. Structure visualization and analysis of HP1β were performed by using Missense 3D^6^. A tertiary structure AlphaFold model was used for the structural prediction of HP1β (UniProt ID P83916). Protein sequence conservation analysis was performed by using Clustal W^7^.

### Cellular experiments

The HEK293T cell culture and CBX1 cDNA vector mutagenesis was performed as previously described^1^. Immunofluorescence (IF) staining was performed as previously described^1^. Antibodies used for IF stainings were mouse primary antibody against FLAG (Sigma-Aldrich, F1804) and Alexa Fluor 488 donkey anti-mouse IgG (Life Technologies, A11001). Chromatin separation was performed as previously described^8^. Immunoblotting was performed as previously described^1^. Antibodies used for immunoblotting include mouse monoclonal anti-FLAG (1:1000, Sigma-Aldrich, F3165), histone H3 (1:1000, Cell Signaling, 9715S), α-tubulin (1:7000, Sigma-Aldrich, T6074), Goat Anti-Mouse IgG H&L (HRP) (1:1000, Abcam, ab6789), and Goat Anti-Rabbit IgG H&L (HRP) (1:1000, Abcam, ab6721). Detailed methods can be found in the **Supplementary Document**.

## Clinical Summary

Shared clinical features among the three subjects included developmental delay, autistic features, and hypotonia (**Table 1**); however, the individual with *CBX1* p.C60G variant primarily exhibited speech delay without motor developmental delay. *De novo CBX1* variants [c.119T>C; p.(Leu40Pro), c.178T>G; p.(Cys60Gly), and c.178T>C; p.(Cys60Arg)] were identified through trio-based exome or genome sequencing, and all variants were located within the CD of HP1β (**Fig 1A**). The clinical characteristics of these subjects are summarized in **Table 1**

**Figure 1:**
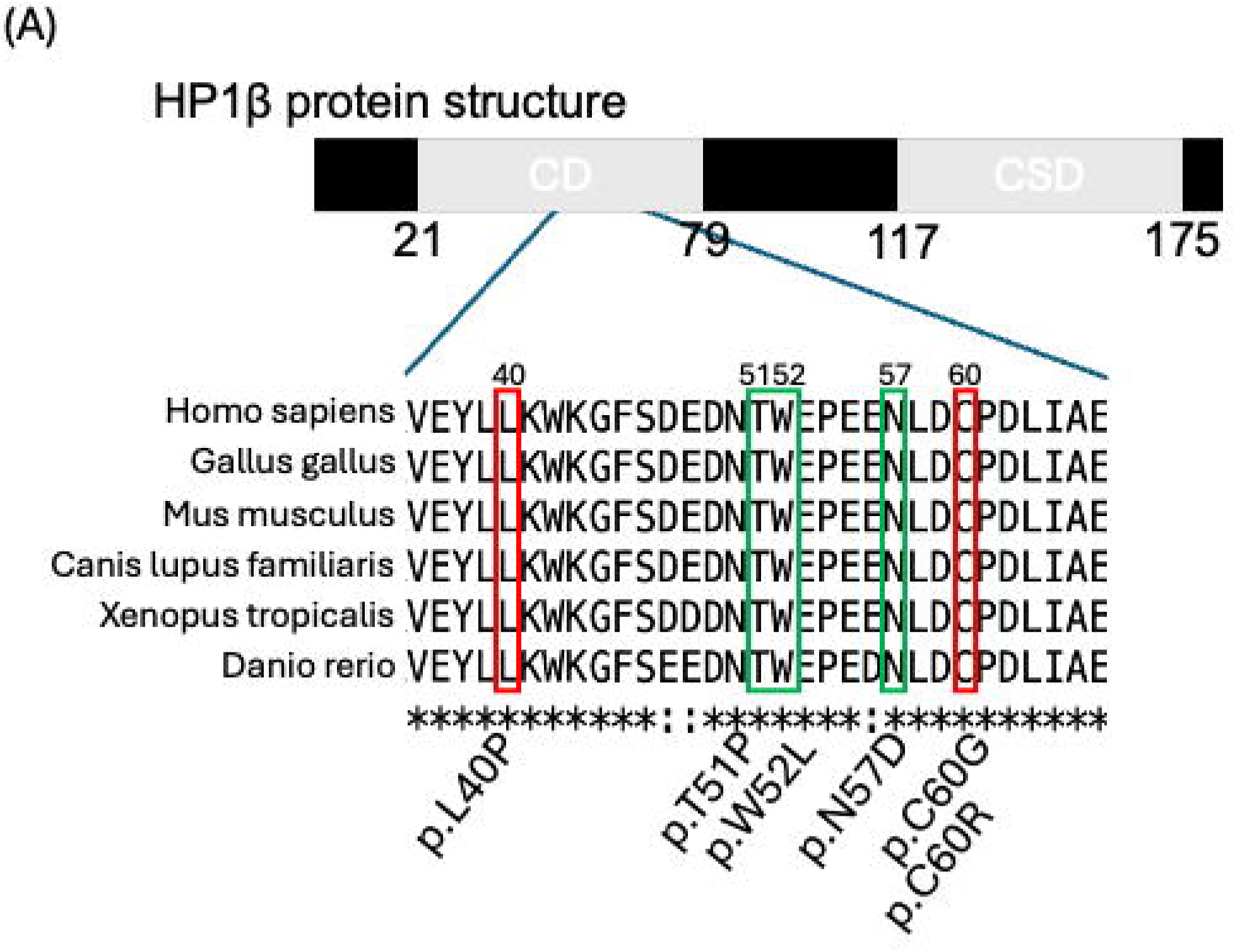
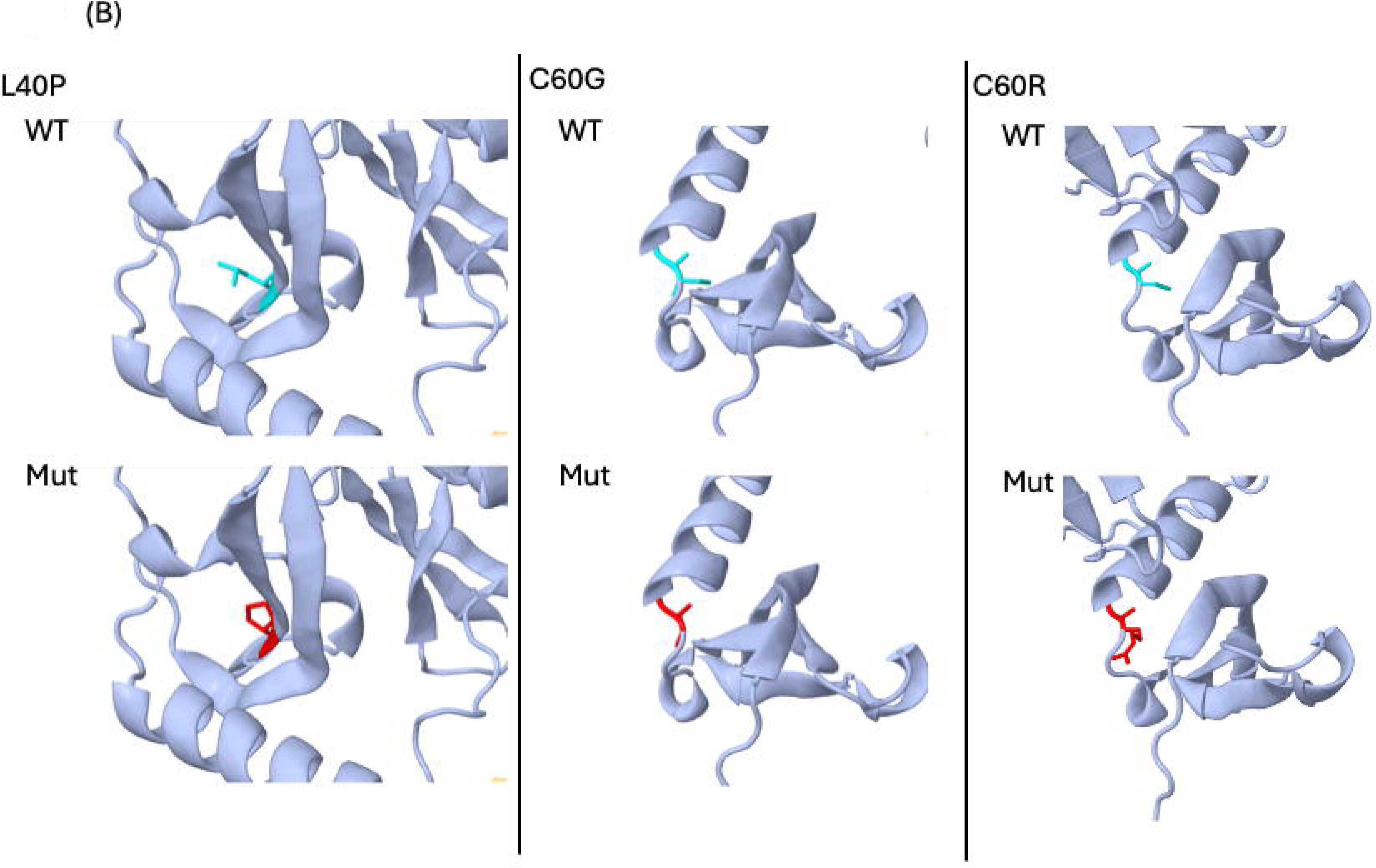
Novel *CBX1* variants. **(A)** Location of the *CBX1* variants. HP1β protein structure is depicted. Amino acids mutated in the previous and current report were marked by green and red rectangle, respectively. CD: chromodomain. CSD: chromoshadow domain. **(B)** 3D protein structural modeling of the mutant HP1β. Left: p.L40P variant was predicted to be damaging due to the introduction of a buried proline. Middle and Right: p.C60G and p.C60R variants were not predicted to cause major structural disruption due to the lack of major structural changes. Wild type amino acids were depicted by turquoise, and mutant amino acids were depicted by red.

**Table 1:** Clinical features of probands with novel *CBX1* variants.

|  | **Proband 1** | **Proband 2** | **Proband 3** | **Individual 1**<br>**(Kuroda et al)** | **Individual 2**<br>**(Kuroda et al)** | **Individual 3**<br>**(Kuroda et al)** |
| --- | --- | --- | --- | --- | --- | --- |
| Sex | Female | Male | Male | Female | Male | Female |
| ***CBX1* variant** |  |  |  |  |  |  |
| cDNA change | NM_001127228.2:<br>c.119T>C | NM_001127228.2:<br>c.178T>G | NM_001127228.2:<br>c.178T>C | NM_001127228.2:<br>c.169A>G | NM_001127228.2:<br>c.155G>T | NM_001127228.2:<br>c.151A>C |
| Protein<br>change | NP_001120700.1:<br>p.(Leu40Pro) | NP_001120700.1:<br>p.(Cys60Gly) | NP_001120700.1:<br>p.(Cys60Arg) | NP_001120700.1:<br>p.(Asn57Asp) | NP_001120700.1:<br>p.(Trp52Leu) | NP_001120700.1:<br>p.(Thr51Pro) |
| Genome<br>coordinate<br>(GRCh38) | Chr17:g.48076886<br>A>G | Chr17:g.48076141<br>A>C | Chr17:g.48076141<br>A>G | Chr17:g<br>48076150T>C | Chr17:g<br>48076164C>A | Chr17:g<br>48076168T>G |
| Method of<br>identification | Trio exome | Trio genome | Trio exome | Trio exome | Trio exome | Trio exome |
| *De*<br>*novo*/inherited | *De novo* | *De novo* | *De novo* | *De novo* | *De novo* | *De novo* |
| **Facial dysmorphisms** | Recessed midface, epicanthal folds, prognathism | Large forehead, small mouth, thin eyebrows | Not reported | Large forehead, frontal bossing, midface hypoplasia, round face, epicanthal folds, upslanting palpebral fissures, flat nasal bridge, short nose, tented upper lip | Large forehead, tall skull with flat occiput, arched eyebrows, small overfolded ears | Large forehead, frontal bossing, round face, almond-shaped eyes |
| **Neurological findings** |  |  |  |  |  |  |
| Muscle tone | Hypotonia | Right hand hypotonia | Generalized hypotonia | Hypotonia | Hypotonia | Hypotonia |
| Brain anomaly | No | No | Mild cortical and subcortical atrophy | No | Increased extra-axial cerebrospinal fluid and mild ventricular dilatation | No |
| Seizure/Epilepsy | No | No | No | No | No | No |
| Other | Unsteady ataxic gait and tires easily |  | Motor and vocal tics | Clumsiness | Joint hypermobility | Mild joint hypermobility |
| **Neurodevelopmental** |  |  |  |  |  |  |

| Overall development | Global developmental delays | Primarily speech delay | Severe global developmental delays | Mild-to-moderate global developmental delay | Mild-to-moderate global developmental delay | Global developmental delay |
| --- | --- | --- | --- | --- | --- | --- |
| Motor abilities | Delayed | Dyspraxia | Delayed motor development | Delayed; still clumsy | Delayed | Marked initial motor delay; later improved strength |
| Cognitive abilities | Impaired | Dysgraphia, attention deficit disorder and concentration disorder | Impaired | Mild to moderate intellectual disability | Mild to borderline intellectual disability | Moderate-severe intellectual disability |
| Behavioral issues | Yes | No | Self-injurious behaviors, behavioral dysregulation | Self-stimulation, sensory-seeking behaviors, repetitive hand movements, head banging, and obsessive-compulsive movements | Not reported | Disruptive food-seeking behavior, self-injurious behavior |
| Autism/autistic features | Moderately severe autism spectrum disorder | Stereotypical behaviors, echolalia, restricted activities | Severe autism spectrum disorder, stereotypies, restricted interests | Autism spectrum disorder, | Autism spectrum disorder, | Severe autism spectrum disorder |
| Verbal abilities | Non-verbal | Present | Non-verbal | Non-verbal | Expressive language delay | Non-verbal |
| Age at walking independently | Between 2 years and 3 years | Between 1 year and 2 years | Not achieved | Between 1 year and 2 years | Between 2 years and 3 years | Between 3 years and 5 years |
| Age with first words | Non-verbal | Between 2 years and 4 years | Non-verbal | Non-verbal | Between 2 years and 3 years | Non-verbal |

## Results

### Computational predictions of variant pathogenicity

Functional predictions from various computational programs were obtained from the Franklin website (**Supplemental Table 1**). The consensus among these programs indicated that all three variants were predicted to be deleterious. These missense variants lead to alterations in amino acids that have been highly conserved throughout evolutionary history (**Fig 1A**). According to Missense 3D, the p.L40P variant was predicted to cause structural damage due to the introduction of a buried proline, which can potentially disrupt protein interactions or compromise protein stability^9,10,11^. In contrast, no structural damage was predicted for two other variants (**Fig 1B**).

### In vitro functional assays

To evaluate the effects of the novel *CBX1* variants, we examined the cellular phenotypes using a FLAG-*CBX1* expression vector in HEK293T cells. The intracellular distribution of HP1β was evaluated using immunofluorescence (IF) staining with an anti-FLAG tag antibody. Consistent with previous findings^1^, IF staining of WT HP1β revealed distinct punctate foci (**Fig 2A**). In contrast, the FLAG-HP1β variants harboring the p.L40P and p.C60R did not exhibit these foci, instead showed uniform nuclear staining, signifying the mislocalization of the mutant HP1β protein (**Fig 2A**). The FLAG-HP1β with the p.C60G mutation exhibited smaller and less distinct foci, with a predominant pattern of uniform nuclear staining, indicating an intermediate phenotype between WT and p.L40P/p.C60R (**Fig 2A**).

**Figure 2:**
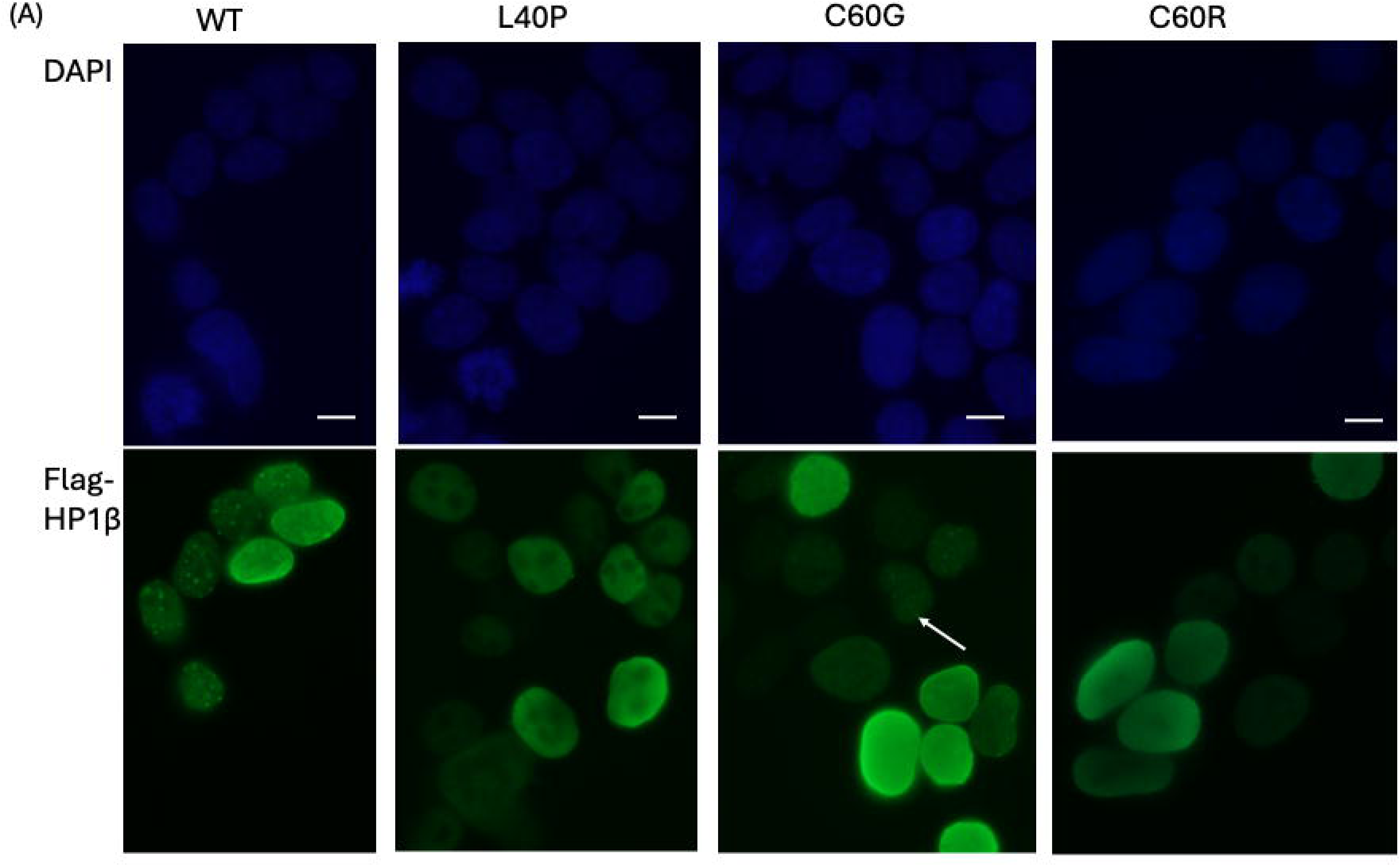
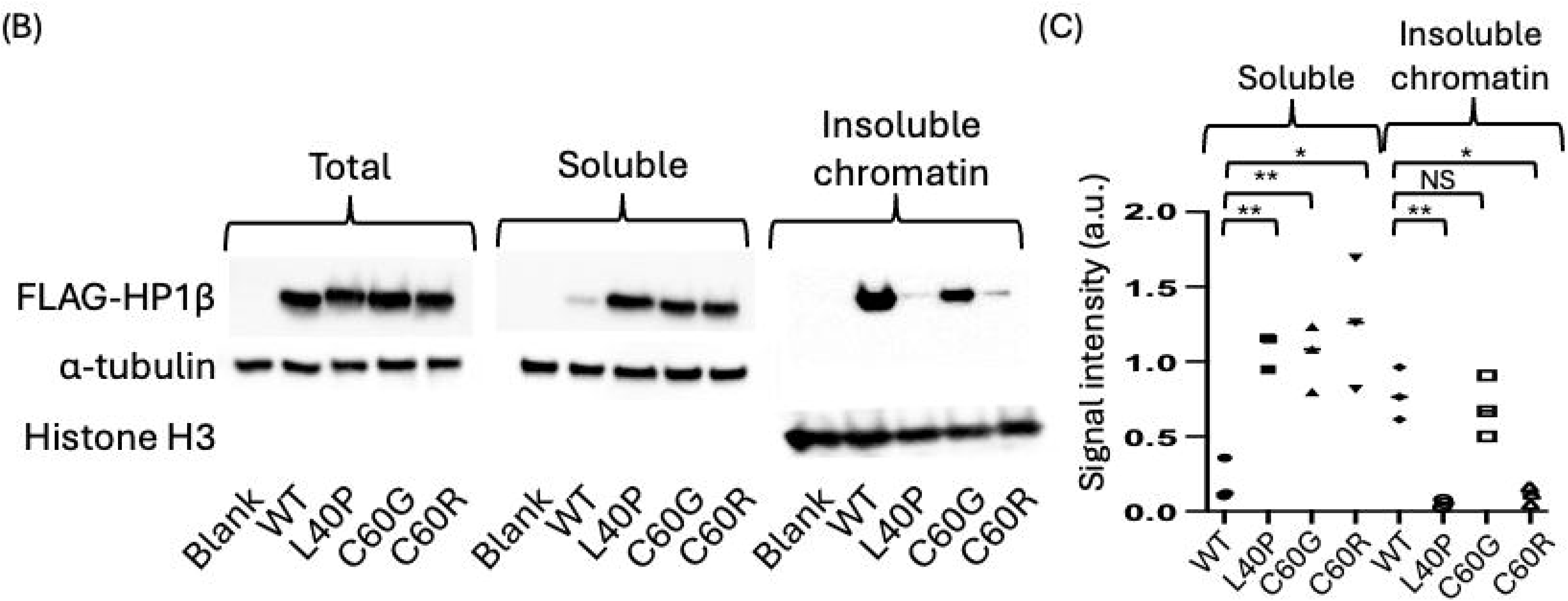
Intranuclear distribution alteration of mutant HP1β. **(A):** Immunofluorescent staining of FLAG-HP1β. FLAG staining of the WT vector revealed that FLAG-HP1β forms well-demarcated punctate foci, however, p.L40P, p.C60G, and p.C60R mutant vectors showed homogenous nuclear staining, although p.C60G mutant vector demonstrated smaller less-demarcated punctate foci indicated by an arrow. Top: DAPI staining. Bottom: FLAG staining. Scale bars are 5 μm. **(B)** Immunoblotting of subcellular fractionation lysates. p.L40P, p.C60G, and p.C60R mutant FLAG-HP1β demonstrated reduced abundance compared to the WT FLAG-HP1β in the insoluble chromatin fraction. Meanwhile p.L40P, p.C60G, and p.C60R mutant FLAG-HP1β demonstrated increased abundance in the soluble fraction. α-tubulin and histone H3 are loading controls. **(C)** Signal intensity measurement of immunoblotting. Signal intensities of FLAG-HP1β bands were normalized against those of α-tubulin (soluble fraction) or histone H3 (insoluble chromatin fraction) and those of the corresponding total cellular fraction. Data represent three independent replicates. Statistical significance was determined using Student’s t-test. *: p < 0.05; **: p < 0.01; NS: not significant.

Given that pathogenic *CBX1* variants result in compromised HP1β-H3K9me3-marked chromatin binding, we hypothesized that the novel HP1β mutants would demonstrate reduced chromatin-binding affinity. Accordingly, WT and mutant FLAG-*CBX1* cDNA vectors were used for immunoblotting of lysates fractionated into soluble and insoluble chromatin components (**Fig 2B**). Effective separation of chromatin was validated by the presence of histone H3 and the absence of α-tubulin in the chromatin fraction. Although the total cellular lysates had comparable levels of FLAG-HP1β, the chromatin fraction had significantly reduced levels of mutant FLAG-HP1β relative to WT. In contrast, the soluble fraction had a markedly higher abundance of mutant FLAG-HP1β than that of WT, indicating an altered intracellular distribution of mutant FLAG-HP1β (**Fig 2B**). Notably, the chromatin fraction had a lower abundance of p.L40P and p.C60R mutant FLAG-HP1β than that of p.C60G mutant FLAG-HP1β, suggesting that p.L40P and p.C60R mutant HP1β had a weaker chromatin-binding capacity than p.C60G (**Fig 2B**). This finding aligns with the IF staining results, suggesting that the p.C60G mutant FLAG-HP1β demonstrates a phenotype that is intermediate between the p.L40P/p.C60R mutant and WT.

## Discussion

In this report, we present three cases of developmental disabilities with novel *de novo CBX1* variants. The clinical features in children with *CBX1* variants typically include global developmental delay, intellectual disability, hypotonia, and autistic features. Notably, Proband 2 primarily exhibited speech delay without concurrent motor developmental delay. Importantly, he has an overall milder presentation and does not have intellectual disability or severe autism. These findings imply that *CBX1*-related syndrome may lead to milder neurodevelopmental symptoms without necessarily affecting motor development. Further clinical studies are warranted to delineate the spectrum of neurodevelopmental phenotype of the *CBX1*-related syndrome.

Interestingly, the Proband 2 who manifested with milder developmental delay was found to have a p.C60G variant, which demonstrated an intermediate cellular phenotype between the WT and the p.L40P/p.C60R variants. This observation may indicate a possible genotype-phenotype correlation. Although this observation is based on a single instance, the extent of chromatin binding by mutant HP1β may serve as a predictor of neurodevelopmental outcomes.

At the molecular level, this study elucidated that variants associated with *CBX1*-related syndromes lead to a generalized reduction in the abundance of HP1β in chromatin. This observation underscores the essential role of the trimethylated histone H3K9-HP1β interaction in tethering HP1β to chromatin. Although HP1β primarily targets H3K9me3-marked heterochromatin, HP1β also exists in euchromatin^12,13^. Consequently, the global decrease in HP1β in chromatin suggests that the effects of *CBX1* variants extend beyond the genomic regions marked by H3K9me3. Further research is warranted to unravel the molecular mechanisms underlying *CBX1*-related syndrome.

Currently, only missense variants within CD of *CBX1* are associated with a human disease^1^. HP1β encoded by *CBX1* forms dimer with HP1 proteins and any dimers incorporating mutant HP1β are expected to be dysfunctional. Based on this model, we proposed dominant-negative effects of mutant HP1β in this *CBX1*-related syndrome^1^. A remaining question includes whether *CBX1* missense variants outside of CD or loss-of-function variants are associated with human disorders. In gnomAD, missense variants’ z-score was 3.25, suggesting significant constraint against missense variation^14^. Missense Tolerance Ratio (MTR) viewer indicates that amino acid sequences comprising CD are particularly intolerant to missense variants, but there is a C-terminus chromoshadown domain, another functional domain of HP1β, demonstrating modest intolerance^15^(**Supplemental Fig 1**). In gnomAD, LOEUF (“loss-of-function observed/expected upper bound fraction”) score was 0.676 with the pLI of 0.4, suggesting that *CBX1* is somehow constrained against loss-of-function variants in human^14^. Further studies are warranted to investigate whether *CBX1* missense variants outside of CD or loss-of-function variants are associated with human disorders.

In conclusion, this report provides additional evidence that missense variants within the CD of HP1β cause a syndromic neurodevelopmental disorder, *CBX1*-related syndrome. To date, this syndrome has been exclusively linked to missense variants of *CBX1*. However, these missense variants present a significant challenge in clinical diagnostics because of the difficulty in predicting their functional implications. It is clinically pertinent to further delineate the variant spectrum associated with *CBX1*-related syndrome to facilitate the identification of affected individuals.

## Data Availability

All data produced in the present study are available upon reasonable request to the authors.

## Acknowledgements

We thank the patients with *CBX1* variants and their families who participated in this study.

---

**Supplementary Figure 1:** Missense Tolerance Ratio (MTR) of HP1β. Top HP1β protein structure. CD: chromodomain. CSD: chromoshadow domain. X-axis: HP1β protein position. Y axis: MTR score. Low MTR score suggests missense constraint. Horizontal lines show gene-specific MTR percentiles 5^th^ (Green), 25^th^ (Orange), 50th, and neutrality (Blue) (MTR = 1.0).

