## Supplemental Table 1 for "Expanding clinical and variant spectrum of *CBX1*-related syndrome: Report of three novel cases"

**Supplemental Table 1: Computational predictions of the effects of the novel *CBX1* variants.**

|  | p.Leu40Pro | p.Cys60Gly | p.Cys60Arg |
| --- | --- | --- | --- |
| Franklin | VUS | VUS | VUS |
| Aggregated Prediction | Deleterious (0.87) | Deleterious (0.86) | Deleterious (0.86) |
| Revel | Deleterious (Moderate) (0.9) | Deleterious (Moderate) (0.87) | Deleterious (Moderate) (0.86) |
| AlphaMissense | Deleterious (Strong) (1) | Deleterious (Strong) (0.999) | Deleterious (Strong) (1) |
| MutationAssessor | Hi (3.98) | Hi (3.75) | Hi (4.17) |
| SIFT | Deleterious (Supporting) (0) | Deleterious (Supporting) (0) | Deleterious (Supporting) (0) |
| MT | Deleterious (1) | Deleterious (1) | Deleterious (1) |
| FATHMM | Uncertain (-0.64) | Uncertain (-0.79) | Uncertain (-0.69) |
| DANN | Deleterious (1) | Deleterious (0.99) | Deleterious (1) |
| MetaLR | Deleterious (low) (0.68) | Deleterious (low) (0.69) | Deleterious (low) (0.69) |
| PrimateAI | Deleterious (Moderate) (0.91) | Deleterious (Supporting) (0.81) | Deleterious (Moderate) (0.88) |
| BayesDel | Deleterious (Moderate) (0.37) | Deleterious (Moderate) (0.36) | Deleterious (Moderate) (0.35) |
| Mutation TasterPrediction | Disease causing | Disease causing | Disease causing |
|  | amino acid sequence changed | amino acid sequence changed | amino acid sequence changed |
|  | protein features (might be) affected | protein features (might be) affected | protein features (might be) affected |
