## Supplemental Document for "Expanding clinical and variant spectrum of *CBX1*-related syndrome: Report of three novel cases"

**Supplementary Document**

**Material and Method:**

*CBX1* variant computational prediction:

In silico pathogenicity prediction was performed by using Franklin (https://franklin.genoox.com/clinical-db/home) and MutationTaster^5^. Structure visualization and analysis of HP1β were performed by using Missense 3D^6^. A tertiary structure AlphaFold model was used for the structural prediction of HP1β (UniProt ID P83916). Protein sequence conservation analysis was performed by using Clustal W^7^. Protein bank ID used for the sequence conservation analysis were P83916 (Homo sapiens), P83917 (Mus musculus), A0A8I3NPU6 (Canis lupus familiaris), A0A8V0ZR83 (Gallus gallus), Q28HD9 (Xenopus tropicalis), and F1QM86 (Danio rerio).

Cellular experiments:

The HEK293T cell line was cultured in DMEM with D-Glucose and sodium pyruvate (Life Technologies, 11360070) supplemented with 10% FBS and 0.2% penicillin-streptomycin. All cells were cultured at 37°C in 5% CO2. The Myc-DDK-tagged-*CBX1* expression vector was purchased from Origene (RC205672). *CBX1* variants were introduced using the Q5 Site-Directed Mutagenesis Kit (New England Biolabs Inc., E0554S) following the manufacturer’s protocol. Sanger sequencing confirmed the introduced variants and ruled out additional, nonspecific changes.

Immunofluorescence (IF) staining:

HEK293T cells were transfected with 0.5µg of *CBX1* expression vectors (wild type (WT), p.L40P, p.C60G, or p.C60R cDNA) using Lipofectamine 2000 (Life Technologies, 11668-030). Cells were settled on poly-D lysine coated coverslips. After 24 hours of transfection, cells were fixed in 4% formaldehyde in PBS for 10 min at room temperature (RT), permeabilized in 0.5% Triton X-100 in PBS for 5 min at RT, then incubated with IF blocking buffer (2% FBS, 2% BSA, 0.1% Tween, 0.02% Sodium Azide in PBS) for 20 min at RT. Cells were incubated with a mouse primary antibody against FLAG (1:500, Sigma, F1804) diluted in IF blocking buffer for 1 hour at RT, followed by three washes in 0.1% Tween in PBS. Subsequently, cells were incubated in Alexa Fluor 488 donkey anti-mouse IgG (1:1000) diluted in IF blocking buffer for 1 hour at RT. After several washes, cells were mounted on to the slide in Vectashield containing 4′,6-diamino-2-phenilinodole (DAPI, Vector Laboratories, H-1200). Images were captured on an Olympus BX41 wide-field fluorescence microscope, using the Olympus DP73 imaging camera.

Chromatin separation:

HEK293T cells were transfected with 1µg of *CBX1* expression vectors (p.L40P, p.C60G, or p.C60R cDNA) using Lipofectamine 2000 (Life Technologies, 11668-030). Transfected cells were dissociated by using 0.25% Trypsin-EDTA (Life Technologies, 25200056). Cells were then washed with PBS once and cells were lysed with lysis buffer (20 mM HEPES, 10 mM KCl, 100 mM NaCl, 1.5 mM MgCl_2_, 0.34 M sucrose, 10% glycerol, 10 mM sodium fluoride, 10 mM β-glycerophosphate, 10 mM sodium butyrate, 1 M DTT, 20% Triton X and complete protease inhibitor cocktail (Roche)). To obtain soluble cell extracts, cells were lysed with lysis buffer and centrifuged at 1,500g at RT. Then, a portion of supernatant was harvested as soluble fraction cellular lysate. DNA in the chromatin fraction was digested by treatment with benzonase (Novagen, Merck Millipore) and the lysates were harvested as insoluble chromatin fraction cellular lysate.

Immunoblotting:

Lysates were mixed with 4x sample buffer (4x Laemmli buffer, Biorad, 1610747, with 10% 2-mercaptoethanol, Biorad, 1610710). Then, samples were denatured at 96 degrees for 5 min. Protein lysates were then subjected to standard sodium dodecyl sulphate-polyacrylamide gel electrophoresis (SDS-PAGE) using 4-15% polyacrylamide Tris/lycine gels (Bio-Rad, 4561086). Proteins were blotted to PVDF LF membrane (Bio-Rad, 1620264) using Biorad’s Trans-Blot Turbo system (Bio-Rad, 1704150). After transfer, the PVDF membrane was blocked with EveryBlot Blocking Buffer (Biorad, 12010020) at RT, and subsequently, blotted membrane was incubated with primary antibodies. Primary antibodies used for immunoblotting were monoclonal anti-FLAG (1:1000, Sigma-Aldrich, F3165), histone H3 (1:1000, Cell Signaling, 9715S), and α-tubulin (1:7000, Sigma- Aldrich, T6074). After primary antibody incubation, membrane was washed three times. Then, the membrane was incubated with secondary antibody Goat Anti-Mouse IgG H&L (HRP) (1:1000, Abcam, ab6789) or Goat Anti-Rabbit IgG H&L (HRP) (1:1000, Abcam, ab6721). The membrane was incubated with a 1:1 ratio of Clarity Western ECL Substrate (Biorad, 1705061) and then Chemi-Doc imaging system (Biorad) was used to detect western blot signals. Signal intensities were quantified using ImageJ. FLAG-HP1β signal intensity in the insoluble chromatin and soluble fractions was normalized to histone H3 and α-tubulin, respectively. The resulting values were further normalized to the abundance of FLAG-HP1β, which was normalized against α-tubulin, in the corresponding total cell lysates to correct for differences in FLAG-CBX1 cDNA transfection efficiency among samples. Three independent experiments were performed, and signal intensities from these biological replicates were used for quantitative analysis. Student t-test was used for statistical analysis. Dot plots were generated using GraphPad Prism

All experiments were performed at least in duplicates, and consistent results were observed among replicates.
