## Supplementary figures and images for "Expanding clinical and variant spectrum of *CBX1*-related syndrome: Report of three novel cases"

### Supplemental Fig 1

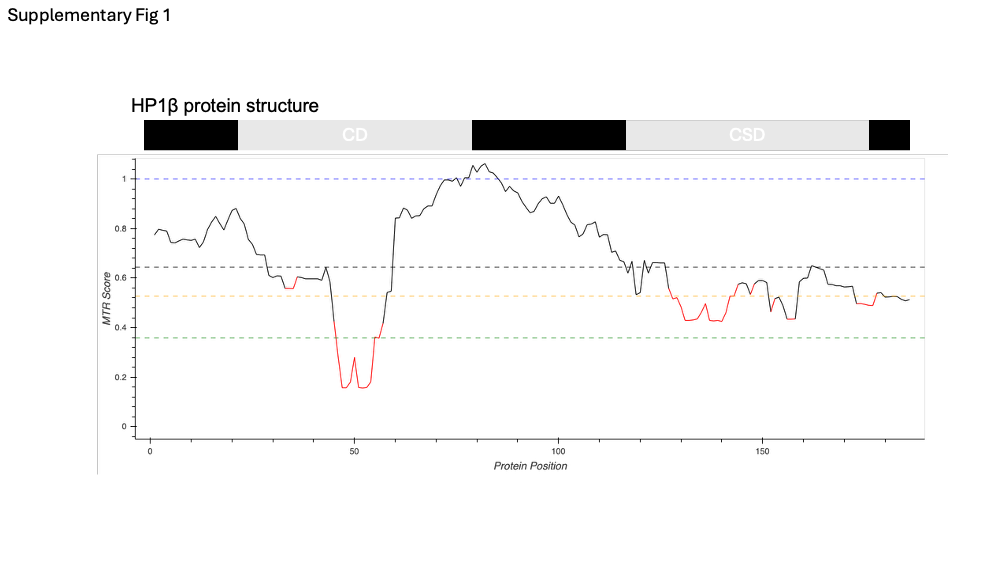
